# Trends and Determinants of changes in HIV Testing Uptake Among Adolescent Girls and Young Women in Mainland Tanzania: a secondary analysis of the 2011/12, 2016/17, and 2022/2023 National Survey Data

**DOI:** 10.64898/2025.12.02.25340958

**Authors:** Deogratius W. Kinoko, Anthony C. Kavindi, Dickson Doto, Grace Mtavangu, Ahmed Y. Nyaki, Sia E. Msuya, Michael J. Mahande

## Abstract

**Background:** Adolescent girls and young women (AGYW) in sub-Saharan Africa, including Tanzania, remain disproportionately vulnerable to HIV. Despite expanded HIV testing services (HTS), many AGYW remain unaware of their HIV status.

**Aim:** This study aimed to assess trends and drivers of changes in HIV Testing Uptake among AGYW in mainland Tanzania using data from three national surveys conducted over time.

**Methodology:** A cross-sectional secondary data analysis was conducted using data from the Tanzania HIV and Malaria Indicator Survey (2011/12) and the Tanzania HIV Impact Surveys (2016/17 and 2022/23). Descriptive statistics summarized AGYW characteristics. A P-trend chi-squared test was used to identify significant changes in HIV testing uptake across survey years. A multivariable Poisson decomposition analysis was used to assess the drivers of changes in HIV testing uptake among AGYW in mainland Tanzania over time, as observed in the THMIS 2011/12, THIS 2016/17, and THIS 2022/23 surveys.

**Results:** Overall HIV testing uptake among AGYW increased from 54% (2011/12) to 61% (2016/17) and 64% (2022/23. Changes in the study participants characteristics in the phase II compared with both Phase III and I contributed 68.5% in increase HIV testing uptake, the main contributors to changes in testing uptake were being aged 20-24 years (39.6%, p-value< 0.001) and women with one sexual partner (38.8%, p-value<:0.001), followed by being with a STI history (24.9%, p-value<:0.001). Whereas 31.5% were attributed to the coefficient.

**Conclusion:** HIV testing uptake among AGYW in Tanzania has improved over time, with significant disparities between adolescents and young women. Being aged 20-24 years and changing sexual behavior patterns emerged as the strongest contributor to the observed changes. These findings highlight the need for age-specific strategies, intensifying adolescent-focused interventions while sustaining efforts among young women and reinforcing integrated reproductive health and HIV services.

## Background

HIV/AIDS remains a major public health challenge globally, with sub-Saharan Africa (SSA) bearing the highest burden(1). In 2023, SSA accounted for 62% of the 670,000 new HIV infections. Alarmingly, each week, nearly 4,000 AGYW acquire HIV globally, and approximately 3,100 of these infections occur in SSA(1). In Tanzania, the epidemic is generalized, and AGYW face significantly higher HIV incidence(0.33%) compared to their male peers(0.00%)(2).

In Nigeria, the trend of HIV testing uptake increased from 33.6% in 2011 to 58.8% in 2016(3). In Lesotho, it rose rapidly from 62.2% to 72.5% in the years 2009/10 and 2014/15, respectively(4), and Tanzania from 53.7% in 2011/12 to 63% in 2022/23(2).

Although national HIV testing services have expanded, testing uptake among AGYW remains suboptimal, with 40.5% of HIV-positive AGYW still unaware of their status in 2022/23, falling short of the first UNAIDS’ 95–95–95 targets(1,2). Previous studies have revealed that approximately 44-66% of new HIV infections are from HIV-infected persons who are unaware of their HIV status(5).

Increasing uptake of HIV testing and counselling and decreasing the number of undiagnosed people is identified as a priority area for HIV prevention(6). Despite ongoing initiatives like PITC, CITC, HIV self-testing, and DREAMS(7,8). Barriers such as stigma, limited youth-friendly services, and age-related legal constraints hinder testing, particularly among adolescents aged 15–19 years(6,9). Shifts in sociodemographic characteristics, HIV prevention strategies, and testing modalities may influence patterns of HIV testing uptake among AGYW.

Therefore, this study aims to determine how these factors are changing with time, and recommendations on what should be done to address the factors that contribute to the uptake of HIV testing among AGYW in mainland Tanzania using secondary data from THMIS (2011/2012) and THIS (2016/2017 and 2022/2023). Also, the study may provide essential information to the MoH and stakeholders that is needed for setting priorities in the allocation of HIV testing services and planning towards focused interventions to address HIV testing uptake among AGYW. Additionally, the findings may contribute to accelerating progress toward the first 95 target of the UNAIDS 95-95-95 goals in Tanzania.

### Conceptual framework

#### Conceptual framework for analysis of factors associated with HIV testing uptake among AGYW in mainland Tanzania

This study adopts Andersen’s Behavioral Model of Health Services Utilization to conceptualize the relationship between various individual and contextual factors and HIV testing uptake among AGYW(10), previously applied in studies examining HIV-related behaviors (11,12). The model comprised individual, societal, and health system levels of analysis for studying health services use and its determinants(10,12).

In this study, the framework is conceptualized based on the predisposing factors, enabling factors, and need (or perceived need) factors for health care utilization. First, the level of HIV testing uptake is independently influenced by predisposing factors, which typically include individual and contextual characteristics. Second, predisposing factors are assumed to influence HIV testing uptake indirectly through enabling factors. Third, predisposing factors may also operate through both enabling and perceived/need factors to either facilitate or hinder individuals from accessing HIV testing services, as illustrated in **Figure 1**.

**Figure 1:**
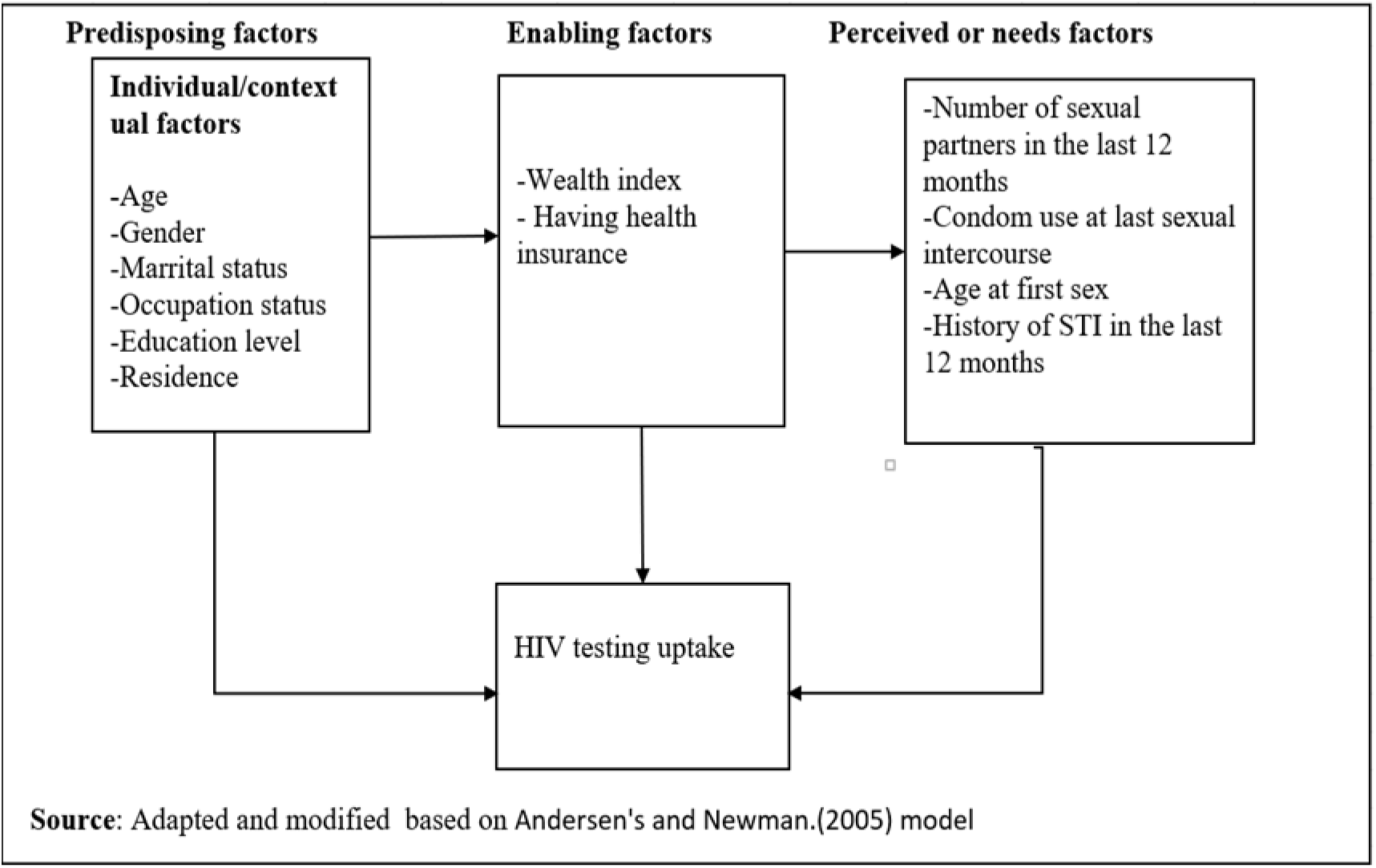
Conceptual framework for analysis of factors associated with HIV testing uptake among AGYW.

## Materials and methods

### Study design and study area

This study uses a cross-sectional design, analyzing data from the Tanzania HIV impact survey conducted between 2011-2012,2016-2017, and 2022-2023. The nationally representative data were collected from all regions of mainland Tanzania. Tanzania, the largest country in East Africa, spans approximately 940,000 square kilometers, including about 60,000 square kilometers of inland water. As of 2022, the estimated AGYW population in mainland Tanzania was 5,988,919(13).

Tanzania HIV/impact surveys implemented by the National Bureau of Statistics (NBS), the Office of the Chief Government Statistician (OCGS) Zanzibar, the National AIDS, STIs and Hepatitis Control Programme (NASHCoP), and the Zanzibar Integrated HIV, Hepatitis, Tuberculosis and Leprosy Program (ZIHHTLP), with technical assistance from the US Centers for Disease Control and Prevention (CDC) and ICAP at Columbia University. The surveys are funded by the US President’s Emergency Plan for AIDS Relief (PEPFAR).

### Sampling design

The Tanzania HIV impact survey used a two-stage stratified cluster sampling design. In the first stage, enumeration areas (EAs) are selected from the national sampling frame using probability proportional to size (PPS), ensuring representativeness by region and urban-rural residence. In the second stage, a systematic sample of households is drawn from each selected EA based on an updated household listing.

### Study population and sample size

The study included all AGYW interviewed in the Tanzania HIV Impact Survey in the years 2011-2012, 2016-2017, and 2022-202. All AGYW from mainland Tanzania interviewed in the Tanzania HIV Impact Survey between 2011/12, 2016/17, and 2022/23 were included in the study. The number of AGYW (15–24) included in the study in the years 2011/12, 2016/17, and 2022/23, after applying weighting, was 4,259, 6,650, and 6,064, respectively. The total weighted sample size of all AGYW included in the study was 16,973 **(Figure 2)**

**Figure 2:**
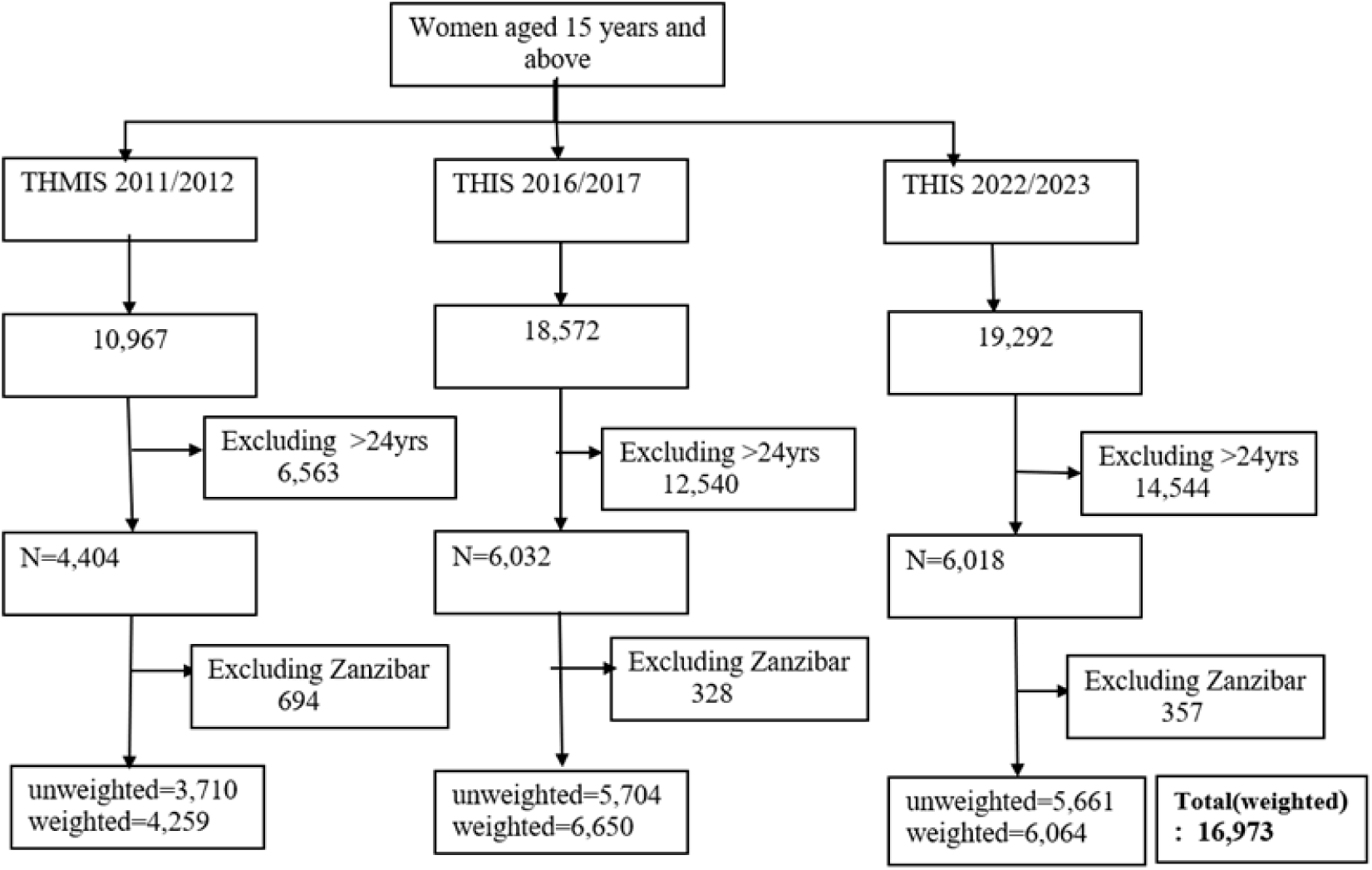
Flow chart for selection of study participants (Adolescent Girls and Young Women)

### Definition of outcome variables

The outcome of interest in this study was ever been tested for HIV and received results, a binary variable if the AGYW reported “Yes” if ever been tested for HIV and received results and “No” if not ever tested for HIV, coded as 1 “Yes” and 0 “No”.

### Independent variables

Independent variables were classified into Predisposing factors, enabling and perceived or needs factors. The predisposing factors includes: age of respondents (0=15-19 and 1=20-24years), place of residence (0=Rural, and 1=Urban), administrative zone (1=Central, 2=Lake, 3=Northen, 4= Eastern, 5=Southern West Highland, 6=Southern Highland, 7= Southern, 8=Western), marital status(0=Never in union, 1=Currently in union, 2=Cohabiting and 3=formerly in union), occupation status(0=Not employed, 1=Employed), education level(0=No education, 1=Primary education and 2=secondary education and above), exposure to TV/radio(0=No and 1=Yes). Enabling factors include: having had health insurance (0=No, and 1=Yes), Wealth index (0=poor,1=middle, and 2=Rich). Perceived or needs factor includes: age at first sex (0=<15, 1=15+), multiple sex partners in the last 12 months (0=No partner, 1=one, 2=two and above), having had STI in the last 12 months (0=No, 1=Yes), condom use in the last sex (0=No and 1=Yes). HIV results from the biomarker test (0=Negative, 1=Positive).

Zones rather than administrative regions were used in order to have consistency across the surveys. Between 2011/12 and 2022/23, some of the regions were divided to form new districts and regions, and thus the 2016/17 and 2022/23 surveys had more administrative regions than the 2011/12 survey. All regions (new and old) belong to the same zones, and the geographical coverage of the zones has remained consistent throughout the surveys. Composition of the administrative regions in their respective zones as follows: Eastern (Morogoro, Pwani, Dar es Salaam); Northern (Kilimanjaro, Tanga, Arusha); Lake (Mwanza, Geita, Mara, Simiyu, Shinyanga, Kagera); Central (Dodoma, Manyara, Singida); Western (Kigoma, Tabora); South West Highlands (Katavi, Rukwa, Mbeya, Songwe); Southern Highlands (Iringa, Njombe, Ruvuma); and Southern (Lindi, Mtwara).

### Data management and analysis

Data cleaning and analysis were performed using STATA version 17.0. The Variables were categorized or recategorized based on previous literature and plausibility. The adult individual and biomarker datasets were merged by unique ID variables, and then appended across survey years to form a pooled dataset for analysis. The data analysis accounted for the complex survey design by incorporating survey weights, primary sampling units (clusters), and strata using the svyset command applied to the defined subpopulation.

Descriptive univariate analyses are shown in frequencies and percentages for categorical variables and continuous variables using mean with respective standard deviation to describe the AGYW characteristics. To assess the trend of HIV testing uptake among AGYW characteristics in Tanzania over time (2011/12,2016/17, and 2022/23), a P trend chi-square was used to identify significant changes in HIV testing uptake across survey years. To determine factors associated with the changes in HIV testing uptake among AGYW, crude and adjusted modified Poisson analysis was performed separately across the survey phases, and the variables that were significant in adjusted modified Poisson analysis was carried to multivariable Poisson analysis. The multivariable Poisson decomposition analysis, an extension of the Blinder and Oaxaca decomposition, was chosen because it is used to identify and quantify the factors contributing to differences in a health outcome between two or more groups or over time(15).

The decomposition analysis was achieved using the “mvdcmp” command. This method allows displaying the contribution of each variable to the overall difference in characteristics or the effect of characteristics. The model partitions the change in HIV testing proportions between the period into two components that are attributed to changes in composition, changes in effects, and the interaction between them(15). Therefore, the study was done in three phases;(phase I between 2011/12 and 2016/17, phase II between 2016/17 and 2022/23, phase III between 2011/12 and 2022/23). The log-binomial decomposition model can be represented as follows:

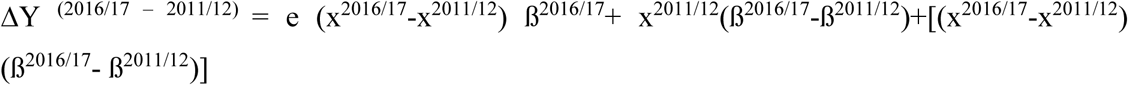

Where:

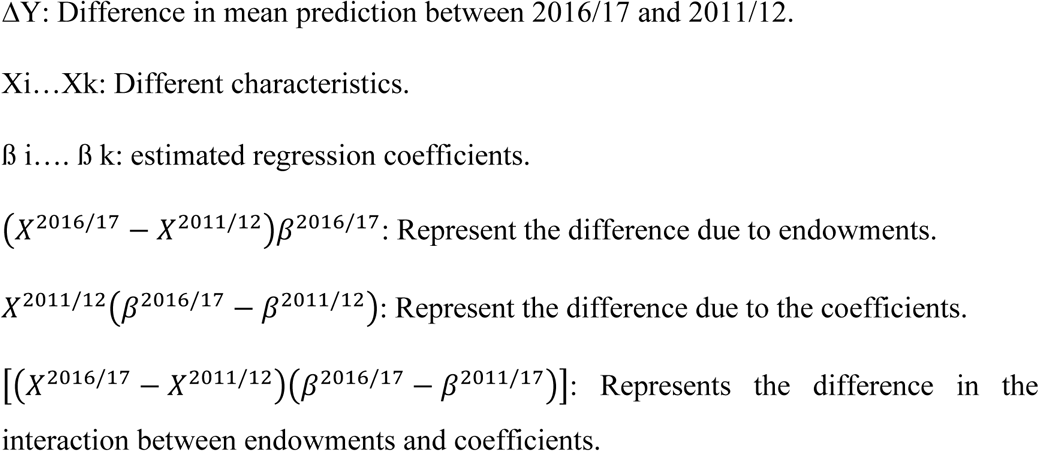

## Results

### Background characteristics of the study participants

A total of 16,973 participants were analyzed across the three surveys, the 2011/12 survey with a mean (SD) age of 19.4(±2.8) (**Figure 2**). Across the three surveys, the majority of participants were aged 15–19 years, although their proportion decreased over time from 56.1% in 2011/12 to 51.4% in 2022/23. The majority of participants were from the Lake zone, with the proportions increasing from 22.3% in THIS 2011/12 to 31.5% in THIS 2022/23. Additionally, the majority resided in rural areas, although this proportion decreased from 73.3% in 2011/12 to 59.3% in 2011/17 and remained relatively stable at 59.4% in 2022/23. Across the three surveys, the majority of AGYW were never married, with the proportion remaining relatively consistent at approximately 56% throughout all survey years. In terms of employment, a majority (67.9%) were employed in 2011/12; however, this number dropped sharply to 25.5% in 2016/17 and slightly rose to 28.7% in 2022/23. Regarding education, most AGYW had attained primary education, although this proportion declined over time from 59.2% in 2011/12 to 48% in 2022/23. Across the three survey years, the majority of AGYW reported having one sexual partner, with proportions increasing from 57.3% in 2011/12 to 63.9% in 2022/23. Additionally, the majority of AGYW reported having had no sexually transmitted infection (STI) in the 12 months before the survey, with the proportions dropping from 96.3% in 2011/12 to 81.3% in 2022/23**(Table 1**).

**Table 1:** Background characteristics of the study participants (weighted) in mainland Tanzania (N=16,973)

| Characteristics | Survey year weighted sample |  |  |
| --- | --- | --- | --- |
|  | 2011/2012<br>(n=4,259)<br>n(%) | 2016/2017<br>(n=6,650)<br>n(%) | 2022/2023 (n=6,064)<br>n(%) |
| **Age(years)** |  |  |  |
| 15-19 | 2,391(56.1) | 3,603(54.2) | 3,115(51.4) |
| 20-24 | 1,868(43.9) | 3,047(45.8) | 2,949(48.6) |
| **Mean (SD)** | 19.1(2.8) | 19.4(2.8) | 19.6(2.8) |
| **Residence** |  |  |  |
| Rural | 3,120(73.3) | 3,943(59.3) | 3,600(59.4) |
| Urban | 1,138(26.7) | 2,707(40.7) | 2,464(40.6) |
| **Zone** |  |  |  |
| Central | 409(9.6) | 698(10.5) | 542(8.9) |
| Lake | 948(22.3) | 1,129(27.5) | 1,928(31.8) |
| Northern | 371(8.7) | 750(11.3) | 640(10.6) |
| Eastern | 686(16.1) | 1,309(19.7) | 1,234(20.4) |
| South West Highland | 694(16.3) | 813(12.2) | 594(9.8) |
| Southern Highland | 442(10.4) | 489(7.4) | 322(5.3) |
| Southern | 452(10.6) | 190(2.9) | 226(3.7) |
| Western | 255(6.0) | 571(8.6) | 577(9.5) |
| **Marital status** |  |  |  |
| Never in union | 2,380(55.9) | 3,705(55.7) | 3,383(55.8) |
| Currently in a union | 1,420(33.4) | 1,875(28.2) | 1,839(30.3) |
| Cohabiting | 268(6.3) | 706(10.6) | 531(8.8) |
| Formerly in union | 191(4.5) | 364(5.5) | 312(5.1) |
| **Occupation status** |  |  |  |
| Not employed | 1,369(32.1) | 4,952(74.5) | 4,323(71.3) |
| Employed | 2,889(67.9) | 1,699(25.5) | 1,741(28.7) |
| **Education Level** |  |  |  |
| No education | 569(13.4) | 551(8.3) | 466(7.7) |
| Primary education | 2,522(59.2) | 3,655(55) | 2,913(48) |
| Secondary/Higher | 1,167(27.4) | 2,445(36.7) | 2,685(44.3) |

<sup>a</sup>-Sex debut 2016/17(n=4,677); <sup>b</sup>-Multiple sexual partners 2016/17(n=6,612);
<sup>c</sup>-Condom use 2011/12(n=2,516),2016/17(n=6,089),2022/23(n=5,804); <sup>e</sup>-Had STI in last 12months 2016/17(n=4,677), 2022/23(n=5,469)

**Table 1 Continued: Background characteristics of the study participants(weighted) in mainland Tanzania (N=16,973)**
| Characteristics | 2011/12(n=4259)<br>n(%) | 2016/17(n=6650)<br>n(%) | 2022/23(n=6064)<br>n(%) |
| --- | --- | --- | --- |
| **Wealth index** |  |  |  |
| Poor | 1,485(34.9) | 2,417(36.3) | 2,472(40.8) |
| Middle | 817(19.2) | 1,389(20.9) | 1,229(20.3) |
| Rich | 1,956(45.9) | 2,845(42.8) | 2,363(38.9) |
| **Exposure to Radio/TV** |  |  |  |
| No | 1,450(34.1) | 2,417(36.3) | 1,842(30.4) |
| Yes | 2,809(65.9) | 4,234(63.7) | 4,222(69.6) |
| **Had health insurance** |  |  |  |
| No | 3,922(92.1) | 5,971(89.8) | 4,817(79.4) |
| Yes | 337(7.9) | 680(10.2) | 1,247(20.6) |
| **Age at first sex <sup>a</sup>** |  |  |  |
| <15 | 1,672(39.3) | 615(13.2) | 351(8.7) |
| 15+ | 2,586(60.7) | 4,062(86.8) | 3,665(91.3) |
| **Multiple sexual partners <sup>b</sup>** |  |  |  |
| No partner | 1,657(38.9) | 583(8.8) | 298(5.0) |
| One | 2,438(57.3) | 3,421(51.7) | 3,642(60.7) |
| Two and above | 164(3.8) | 2,608(39.5) | 2,058(34.3) |
| **Condom use in last sex <sup>c</sup>** |  |  |  |
| No | 1,925(74.2) | 3,197(52.5) | 3,327(57.3) |
| Yes | 668(15.8) | 2892(47.5) | 2,477(42.7) |
| **Had STI in the last 12months<sup>e</sup>** |  |  |  |
| No | 4,099(96.3) | 4,091(87.5) | 4,447(81.3) |
| Yes | 160(3.7) | 586(12.5) | 1,022(18.7) |
| **HIV results from the biomarker test.** |  |  |  |
| Positive | 112(2.6) | 141(2.1) | 78(1.3) |
| Negative | 4,147(97.4) | 6,509(97.9) | 5,986(98.7) |

<sup>a</sup>-Sex debut 2016/17(n=4,677); <sup>b</sup>-Multiple sexual partners 2016/17(n=6,612);
<sup>c</sup>-Condom use 2011/12(n=2,516),2016/17(n=6,089),2022/23(n=5,804); <sup>e</sup>-Had STI in last 12months 2016/17(n=4,677), 2022/23(n=5,469)

### Trends in HIV testing uptake among AGYW characteristics in mainland Tanzania from 2011/12,2016/17, and 2022/23

#### Overall trends in HIV testing uptake

Trends in HIV testing among AGYW (15-24years) who had ever been tested and received results, the proportion increased from 54% in 2011/12 to 61% in 2016/17 and 64% in 2022/23 (**Figure 3**). Adolescent girls aged 15-19 years consistently had lower proportions of HIV testing across three surveys, with the proportions gradually increasing from 35% in THMIS 2011/12 to 40 % in both THIS2016/17 and THIS2022/23. In contrast, young women aged 20-24 years showed significantly higher proportions of HIV testing that increased from 79% in THMIS2011/12 to 86% in THIS2016/17 and reached 90% THIS2022/23 (**Figure 3**).

**Figure 3:**
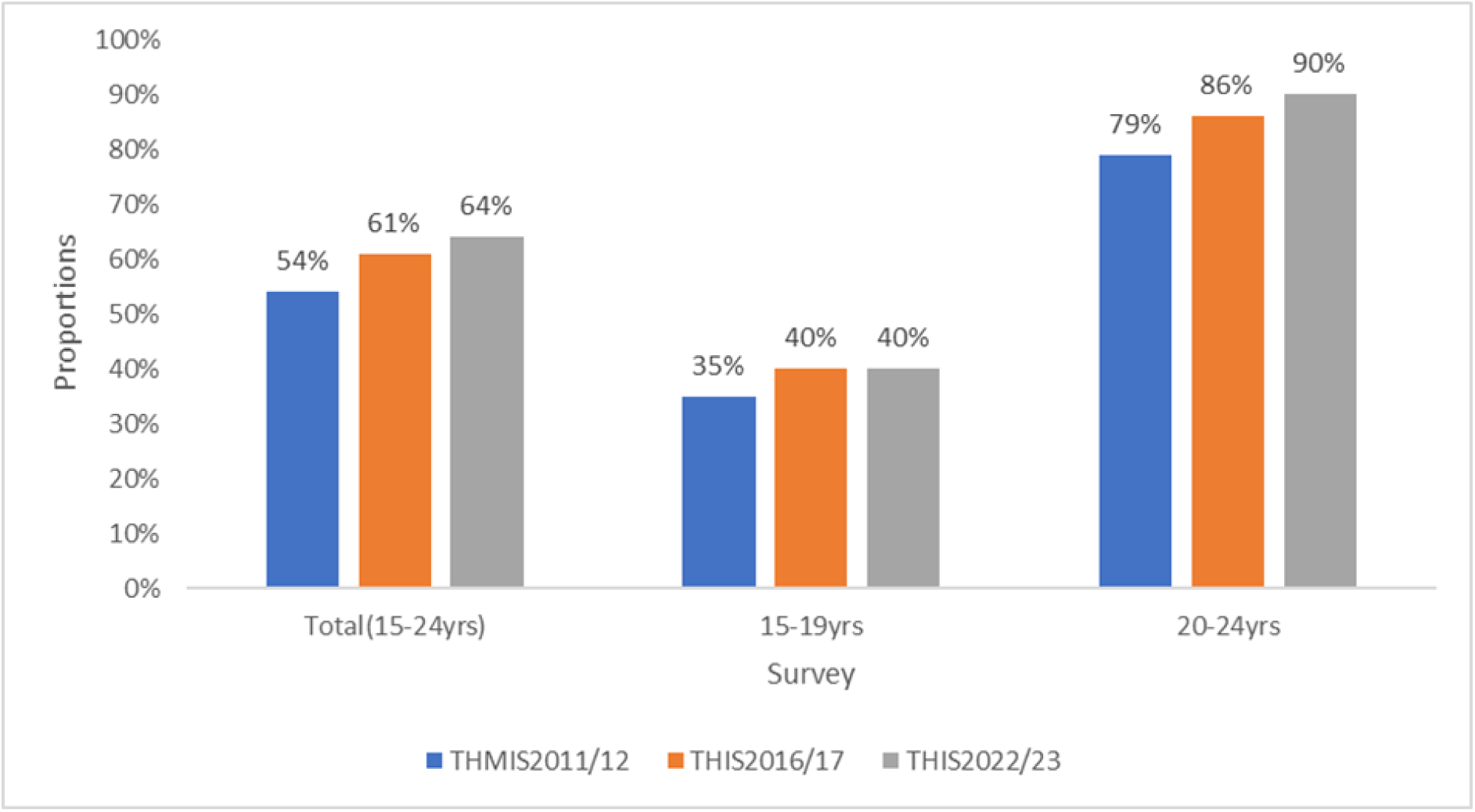
Trends in ever tested for HIV and received results among AGYW in mainland Tanzania, 2011/12, 2016/17, and 2022/23.

#### Factors contributing to changes in HIV testing uptake among AGYW

A multivariable decomposition analysis was used as it provides insight into the sources of difference in the HIV testing uptake proportion of AGYW. They are divided into two components: Characteristic differences(E) and coefficient differences(C).

#### Phase I: 2011/2012 to 2016/2017

During this period, approximately 24% of the increase in HIV testing among AGYW was attributed to changes in population composition (characteristics effect), while 76% was explained by changes in behavior or responsiveness to these characteristics (coefficient effect). Several compositional factors contributed significantly to the observed increase (**Table 2**):

**Table 2:**
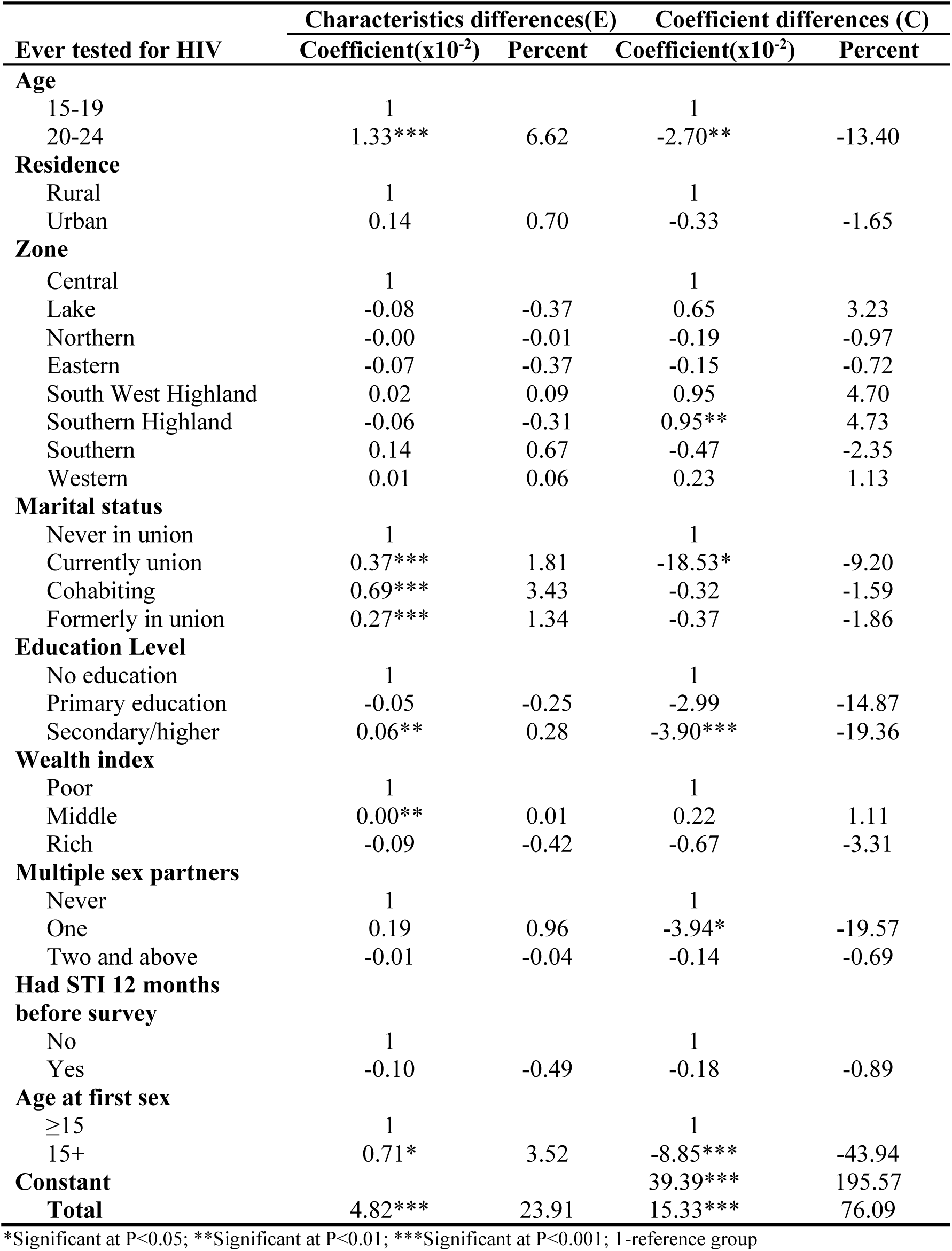
Factors contributing to changes in HIV testing uptake among AGYW in mainland Tanzania in Phase I (N=9,459)

A decrease in the proportion of AGYW aged 20–24 years (**Table 1**) paradoxically contributed (6.6%, p-value< 0.001) to the increase in HIV testing. This suggests increased testing proportion within this age group, despite its shrinking proportion. A reduction in the proportion of AGYW currently in union contributed (1.8%, p-value< 0.001) to the testing increase. This indicates that changes in marital status composition among AGYW had a small contribution but a statistically significant effect on testing trends over the study period. Additionally, a rise in cohabitation rates (**Table 1**) contributed (3.4%, p-value< 0.001) to HIV testing, indicating a notable influence of changing union status on testing trends. Additionally, an increased proportion of age at sexual debut (15+) contributed 3.5% HIV testing among AGYW, reflecting the impact of shifts in sexual behavior patterns on testing uptake (**Table 2**).

The coefficient effects reveal more substantial behavioral changes. Notably, residing in the Southern Highland zone contributed (4.7%, p-value< 0.01), indicating stronger behavioral uptake of HIV testing in that region. The large unexplained variance (constant term = 195.6%) likely reflects external unmeasured influences, such as policy changes (e.g., implementation of youth-friendly services), increased awareness campaigns, or improvements in testing availability (**Table 2**).

#### Phase II: 2016/2017 to 2022/2023

In this phase, the entire increase in HIV testing was attributable to changes in population characteristics (68.5%), while the behavioral component (coefficient effect) accounted for 31.5%. Significant compositional drivers included:

An increase in the proportion of AGYW aged 20–24 (**Table 1**) contributed (39.6%, p-value< 0.001) to the HIV testing proportion, indicating a very strong demographic shift associated with testing trends during the study period. Additionally, an increase in AGYW currently in union proportion contributed an additional (14.3%, p-value<:0.001) of HIV testing proportion, indicating a substantial demographic shift associated with the observed trend. The increase in women reporting one sexual partner added (38.8%, p-value<:0.001) of HIV testing proportion, while a notable increase in proportion of STI history (**Table 1**) reporting accounted (24.9%, p-value<:0.001) of the HIV testing increase, indicating a very strong association with testing trends during the study period (**Table 3).**

**Table 3:**
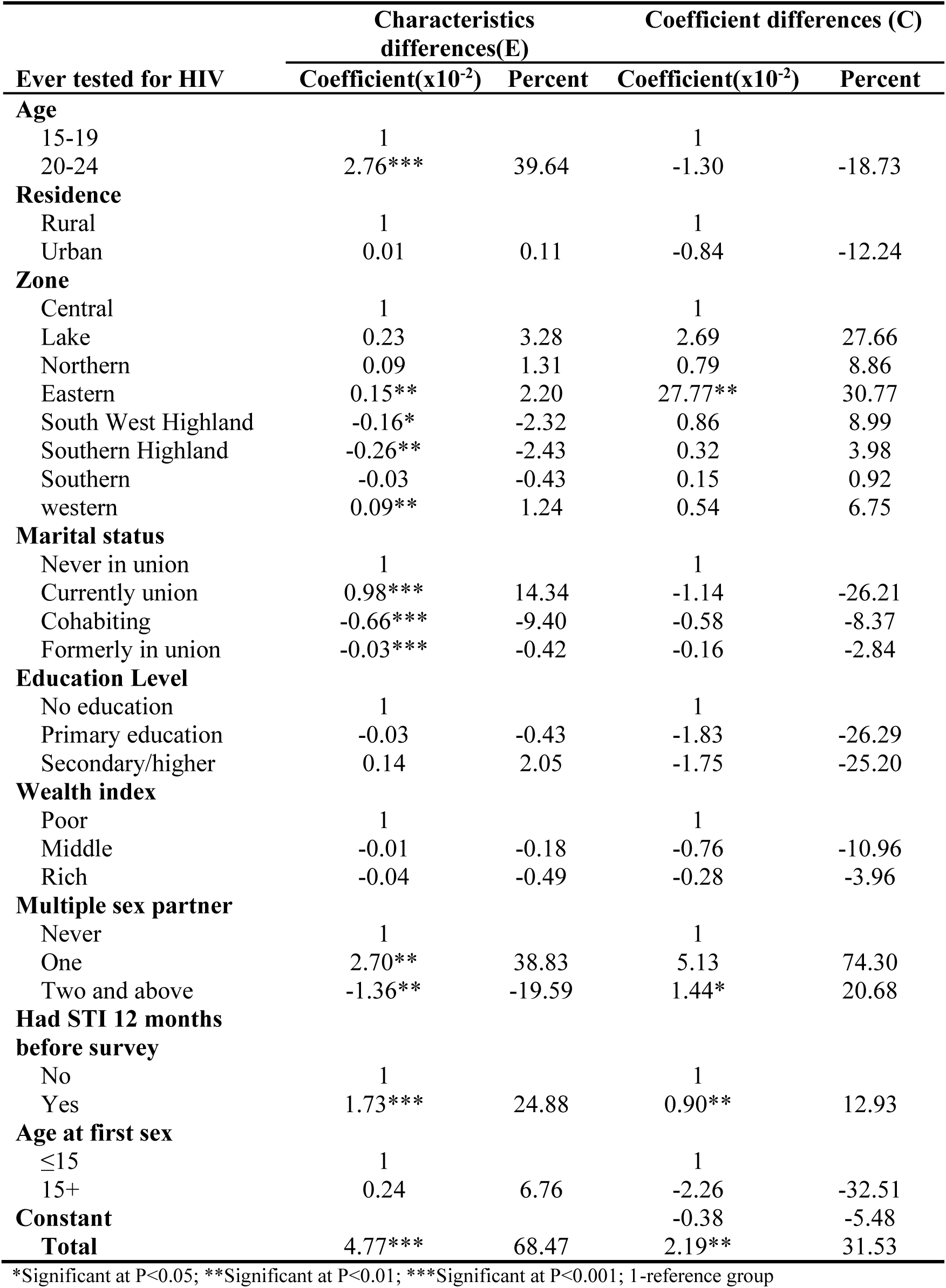
Factors contributing to changes in HIV testing uptake among AGYW in mainland Tanzania in Phase II(N=11,347)

The coefficient effects revealed substantial behavioral changes (31.5%). Notably, residing in the eastern zone contributed 30.8%, indicating stronger behavioral uptake of HIV testing in that region. Having one sexual partner contributed 74.3%; however, was not statistically significant, and having had a history of STI contributed 12.93%, indicating a very strong high-risk perception and how the behavior is associated with an increase in the trend of HIV testing uptake (**Table 3**)

#### Phase III: 2011/12 to 2022/2023

In this phase, population characteristics had a negative contribution to an increase in HIV testing uptake (39.5%), while the behavioral component (coefficient effect) actually had a positive influence (60.5%). Significant compositional drivers included:

An increase in the proportion of young women aged 20-24years (**Table 1**) contributed (11.1%, p-value<:0.001) to an increase in the proportion of HIV testing, representing a notable demographic shift during the study period. Contrarily, a decrease in the proportion of AGYW who were currently in union contributed (3.2%, p-value<:0.001) to an increase in the proportion of HIV testing, indicating that changes in marital status composition influenced testing trends in the opposite direction **(Table 4).**

**Table 4:**
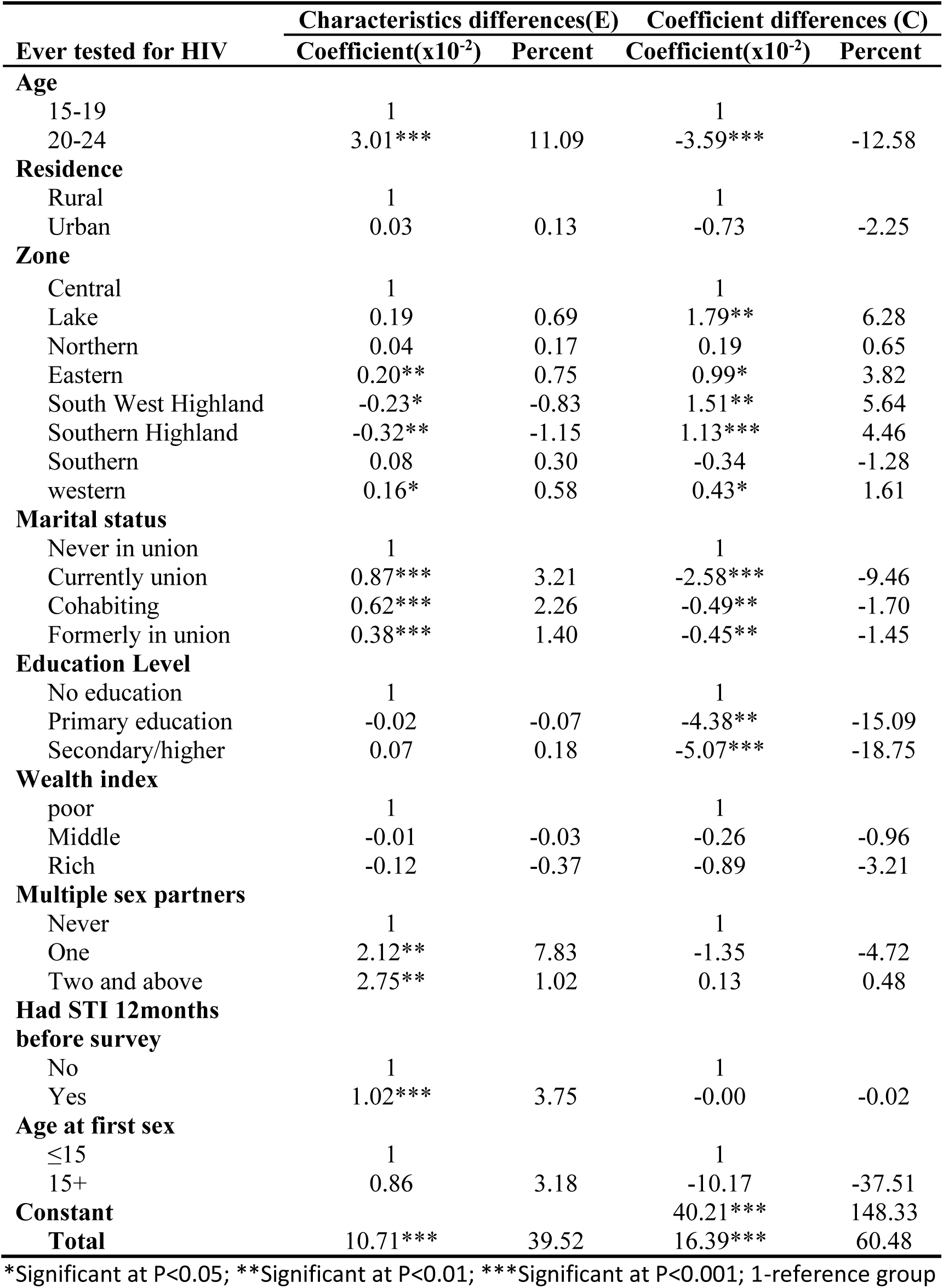
Factors associated with changes in HIV testing proportion among AGYW in mainland Tanzania in Phase III (N=9,308)

| Ever tested for HIV | Characteristics differences(E) |  | Coefficient differences (C) |  |
| --- | --- | --- | --- | --- |
| | Coefficient($\times 10^{-2}$) | Percent | Coefficient($\times 10^{-2}$) | Percent |
| **Age** |  |  |  |  |
| 15-19 | 1 |  | 1 |  |
| 20-24 | 3.01*** | 11.09 | -3.59*** | -12.58 |
| **Residence** |  |  |  |  |
| Rural | 1 |  | 1 |  |
| Urban | 0.03 | 0.13 | -0.73 | -2.25 |
| **Zone** |  |  |  |  |
| Central | 1 |  | 1 |  |
| Lake | 0.19 | 0.69 | 1.79** | 6.28 |
| Northern | 0.04 | 0.17 | 0.19 | 0.65 |
| Eastern | 0.20** | 0.75 | 0.99* | 3.82 |
| South West Highland | -0.23* | -0.83 | 1.51** | 5.64 |
| Southern Highland | -0.32** | -1.15 | 1.13*** | 4.46 |
| Southern | 0.08 | 0.30 | -0.34 | -1.28 |
| western | 0.16* | 0.58 | 0.43* | 1.61 |
| **Marital status** |  |  |  |  |
| Never in union | 1 |  | 1 |  |
| Currently union | 0.87*** | 3.21 | -2.58*** | -9.46 |
| Cohabiting | 0.62*** | 2.26 | -0.49** | -1.70 |
| Formerly in union | 0.38*** | 1.40 | -0.45** | -1.45 |
| **Education Level** |  |  |  |  |
| No education | 1 |  | 1 |  |
| Primary education | -0.02 | -0.07 | -4.38** | -15.09 |
| Secondary/higher | 0.07 | 0.18 | -5.07*** | -18.75 |
| **Wealth index** |  |  |  |  |
| poor | 1 |  | 1 |  |
| Middle | -0.01 | -0.03 | -0.26 | -0.96 |
| Rich | -0.12 | -0.37 | -0.89 | -3.21 |
| **Multiple sex partners** |  |  |  |  |
| Never | 1 |  | 1 |  |
| One | 2.12** | 7.83 | -1.35 | -4.72 |
| Two and above | 2.75** | 1.02 | 0.13 | 0.48 |
| **Had STI 12months before survey** |  |  |  |  |
| No | 1 |  | 1 |  |
| Yes | 1.02*** | 3.75 | -0.00 | -0.02 |
| **Age at first sex** |  |  |  |  |
| $\leq 15$ | 1 | | 1 | |
| 15+ | 0.86 | 3.18 | -10.17 | -37.51 |
| **Constant** |  |  | 40.21*** | 148.33 |
| **Total** | 10.71*** | 39.52 | 16.39*** | 60.48 |

\*Significant at $P < 0.05$ ; \*\*Significant at $P < 0.01$ ; \*\*\*Significant at $P < 0.001$ ; 1-reference group

An increase in the proportion of AGYW with one sexual partner contributed (7.8%, p-value<:0.01) to an increase in HIV testing proportion, indicating a meaningful compositional change in sexual behavior patterns within the population over the study period. Furthermore, an increase in the proportion of AGYW having had STI (**Table 1**) contributed 3.8% to the increase in HIV testing proportion, highlighting its strong influence on the overall trend (**Table 4**).

The coefficient effects revealed more substantial behavioral changes (60.5%). Notably, residing in the lake zone contributed 6.3%, Southern West Highland contributed 5.6%, and Southern Highland contributed 4.5%, indicating stronger behavioral uptake of HIV testing in that region. The large unexplained variance (constant term = 148.3%) likely reflects external unmeasured influences, such as policy changes (e.g., implementation of youth-friendly services), increased awareness campaigns, or improvements in testing availability (**Table 4**)

## Discussion

### Trends of HIV testing uptake

Our findings demonstrate a modest but notable increase in HIV testing uptake among AGYW over the past decade, from 54% in 2011/12 to 64% in 2022/23. Among young women aged 20–24 years, HIV testing coverage improved significantly from 79% in 2011/12 to 90% in 2022/23. In contrast, the uptake among adolescent girls aged 15–19 years increased only slightly, from 35% to 40%, with no substantial progress observed between 2016/17 and 2022/23.

These results are consistent with trends reported in other sub-Saharan African countries. For example, in Lesotho, HIV testing among adolescents increased from 26.5% in 2010/11 to 47.9% in 2015(4), a greater relative improvement than observed in Tanzania. The persistently low uptake among adolescents in Tanzania may reflect structural and policy barriers, such as the requirement for parental consent for individuals under 18 years to access HIV testing services(16). In contrast, countries like Lesotho permit self-consent from age 12, potentially facilitating earlier engagement with HIV services(17). Higher testing rates among young women compared to adolescents may be partially explained by their greater engagement with healthcare services, particularly through antenatal care (ANC) and maternal health programs. Tanzanian national guidelines recommend provider-initiated HIV testing during ANC visits, which may increase testing opportunities for women in this age group.

### Factors contributing to changes in HIV testing

The multivariable Poisson decomposition analysis across all phases revealed, the overall change in HIV testing among AGYW attributed to both changes in population characteristics especially in phase II and III (e.g., increased in; young women aged 20-24 years, currently in union, having had an STI and one sexual partner) and changes in behavioral effects or responsiveness in phase I and III(e.g., improved uptake due to programs or awareness) These factors have also been associated with increase in HIV testing uptake in previous study(22–24)

For example, in phases II and III, the overall change in HIV testing proportions was attributed to changes in population structure by 68.5% and 39.5%, respectively. In all phases, increases in the proportion of AGYW aged 20-24 years, having had STI, being with one sexual partner, and being in union have increased to proportions of HIV testing.

Across all phases, AGYW aged 20–24 years consistently emerged as the most influential factor contributing to the observed increases in HIV testing uptake, highlighting the importance of this age group in HIV response efforts. This finding is similar to a study conducted in Ethiopia. (25). In contrast, earlier evidence from Tanzania (Mahande et al., 2016) identified antenatal care (ANC) visits as the primary driver of testing uptake. The shift in contributing factors may reflect evolving programmatic priorities, including the recent national emphasis on integrated reproductive health and HIV services. Also, it may be partly due to differences in the variables analyzed.

The coefficient effect has contributed 76.1% in phase I and 60.5% in phase III. However, the large unexplained variance (constant term = 195.6% in phase I and 148.3% in phase III) likely reflects external unmeasured influences, such as policy changes (e.g., implementation of youth-friendly services), increased awareness campaigns, or improvements in testing availability.

### Strengths and Limitations of this study

➢ This study comprehensively examines determinants of changes in HIV testing uptake among AGYW, using robust nationally representative survey data spanning over a decade.
➢ The large sample sizes provided sufficient power for decomposition analysis.
➢ The use of secondary data introduces potential issues related to missing or unmeasured variables.
➢ Self-reported HIV testing history may be subject to recall or social desirability bias, particularly among adolescents.
➢ Additionally, certain important contextual and behavioral variables (e.g., stigma, peer influence, knowing a place for HIV testing, alcohol use, and sexual violence) were not captured in the datasets consistently, limiting the interpretation of some observed associations.

## Conclusion

HIV testing uptake among AGYW in mainland Tanzania has improved between 2011/12 and 2022/2023, with notable gains among young women aged 20–24 years. However, progress among adolescents remains limited. The main contributors to changes in testing uptake across all survey periods were being aged 20-24 years and changing sexual behavior patterns, followed by being in a union.

### Recommendations

Integrate HIV testing with reproductive and STI services: Programs should continue strengthening HIV testing during STI care and expand testing access through youth-friendly health services.

Strengthen and promote couple HIV Testing and Counselling (CHTC), particularly for AGYW currently in union or cohabiting relationships. This approach enhances mutual disclosure, reduces stigma, and promotes shared responsibility in HIV prevention and care.

Strengthen school-based sexual and reproductive health (SRH) education: efforts should focus on enhancing their quality, coverage, and effectiveness. The Tanzania Education and Training Policy (ETP) 2014 Edition of 2023 outlines several key including integration of health education in primary, secondary, and higher levels, to overcome health challenges(26).

Conduct qualitative research: Further studies are needed to explore social, cultural, and psychological barriers to HIV testing among adolescents, particularly those aged 15–19 years.

## Data Availability

All data produced are available online at request on the PHIA project website at http://phia.icap.columbia.edu and on the National Bureau of Statistics website at www.nbs.go.tz.

https://www.nbs.go.tz

## List of abbreviations

AGYW: Adolescent Girl and Young Women
CHTC: Couple HIV Testing and Counselling
CITC: Client-Initiated HIV Testing and Counselling
DHS: Demographic Health Survey
HIVST: HIV Self Testing
HTS: HIV Testing Services
MoH: Ministry of Health
NASHCoP: National AIDS, STIs and Hepatitis Control Programme
PITC: Provider-Initiated HIV Testing and Counselling
STIs: Sexually Transmitted Infections
THIS: Tanzania HIV Impact Survey
UNAIDS: United Nations Programme on HIV/AIDS
VCTC: Voluntary Testing and Counselling Center
WHO: World Health Organization

## Declaration

### Ethics Approval and Consent to Participate

Each survey received ethical clearance from the National Institute for Medical Research (NIMR) in Tanzania and the Zanzibar Medical Research and Ethics Committee (ZMREC), as well as approval from relevant international institutional review boards. Informed consent was obtained from all participants before data collection, with assent and parental permission required for minors.

For this secondary analysis, ethical approval to conduct the study was sought from the KCMC University Research and Ethics Review Committee (KURERC) with clearance number PG 180/2024. Permission to access the datasets was granted by the Tanzania National Data Archive (TNADA) and the PHIA project website following submission of a project abstract.

## Availability of Data and Materials

The THIS data and materials used in this study are available for free and on request on the PHIA project website at http://phia.icap.columbia.edu and on the National Bureau of Statistics website at www.nbs.go.tz.

## Acknowledgments

We acknowledge and appreciate the academic staff of the Institute of Public Health–KCMC University for their support. The National Bureau of Statistics for conducting the surveys and providing the dataset.

## Author Contributions

**Conceptualization**: Deogratius W. Kinoko, Sia E. Msuya, Michael J. Mahade, Ahmed Y. Nyaki

**Formal analysis**: Deogratius W. Kinoko, Sia E. Msuya, Michael J. Mahade, Ahmed Y. Nyaki, Anthony C. Kavindi

**Methodology**: Deogratius W. Kinoko, Anthony C. Kavindi, Dickson Doto, Grace Mtavangu

**Supervision**: Sia E. Msuya, Michael J. Mahade, Ahmed Y. Nyaki

**Writing – original draft**: Deogratius W. Kinoko.

**Writing – review & editing**: Sia E. Msuya, Michael J. Mahade, Ahmed Y. Nyaki

